# Pediatric lumbar puncture rates in urban tertiary hospitals in Africa participating in the SURE pediatric tuberculous meningitis treatment trial

**DOI:** 10.64898/2026.11.04.26367254

**Authors:** Constantine Mutata, Christina Zulu, Susan Abarca Salazar, Nancy Nabukeera, Suzanne T Anderson, Trust Mukanganiki, Wendy Mapfumo, Dorothy Murungu, Mario Kanga, Hellen Tukamuhebwa Aanyu, Maria Selinopoulou, Margaret. C. Siu, James A Seddon, Anna Griffiths, Caitlin Muller, Sierra Santana, Elena Frangou, Louise Choo, Angela Crook, Tom Gibb, Diana Gibb, Eric Wobudeya, Sabrina Bakeera Kitaka, Chishala Chabala, Hilda. A. Mujuru, Robin Basu Roy

**Author notes:** **Correspondence:** Robin Basu Roy. These authors contributed equally to this manuscript and are joint first authors (Constantine Mutata, Christina Zulu, Susan Abarca Salazar). Now deceased (Susan Abarca Salazar).

## Abstract

Lumbar puncture (LP) and cerebrospinal fluid (CSF) analysis are essential for the diagnosis and management of central nervous system (CNS) infections, but LP is underutilised in pediatric patients in high burden settings. As part of the pre-screening and screening procedures of the SURE+DP diagnostic sub-study of the SURE pediatric tuberculous meningitis randomised control trial we identified this as a key clinical issue and quantified how often pediatric lumbar puncture was performed at recruiting African hospitals and implemented a package of interventions to increase LP rates.

**Methods:** We evaluated the frequency of pediatric CSF sampling of children aged 28 days to 18 years at four urban tertiary hospitals in Kampala, Lusaka and Harare. We developed a family-facing educational video to support LP consent discussions. In Zambia and Zimbabwe, healthcare workers received training sessions including mannequin-based LP simulation, with knowledge, confidence, experience and perceived barriers assessed before and after training. We assessed the effect of healthcare worker LP training on confidence and LP performance and explored reported barriers to pediatric LP.

**Results:** Median weekly pediatric CSF samples were 1 at both Sally Mugabe and Parirenyatwa hospitals in Harare, 6 at University Teaching Hospital in Lusaka, and 4 at Mulago Hospital in Kampala. Training significantly improved healthcare-worker self-reported confidence to assist (p-value <0.001) and perform pediatric LP (p-value <0.001). At the Zambian site, the proportion of children with coded diagnoses compatible with consideration of LP who had CSF samples increased by 7% in the six months after training compared with the preceding six months (p=0.002). In Zimbabwe, there was an increase in LP activity the week following training, which was not subsequently sustained. Healthcare workers identified parental refusal, limited procedural confidence, delays obtaining consent from male family decision makers, fear of complications, and consumables availability as barriers.

**Discussion:** Pediatric LP was performed infrequently across these tertiary hospitals, and improved healthcare worker confidence following training did not consistently translate into sustained increases in LP performance. Multifaceted sustained approaches combining healthcare worker training with caregiver engagement, system level support and improved resources are needed to improve diagnostic LP use for children with suspected CNS infections.

## Introduction

Tuberculous meningitis (TBM) is an important cause of severe childhood neurological disease in high burden settings and is associated with substantial mortality and neurological disability (1,2). Diagnosis of TBM requires an aseptically collected cerebrospinal fluid (CSF) sample, obtained by lumbar puncture (LP) in the absence of contraindications. This sample is analysed by microscopy, biochemistry, mycobacterial and conventional bacterial culture and rapid nucleic acid amplification tests for TB alongside timely exclusion of alternative diagnoses (3,4). Given it is not possible to reliably clinically discriminate between CNS infections, LP and CSF analysis is the cornerstone of making the correct diagnosis and initiating appropriate treatment for all causes of meningitis. CNS infections affect more than 1.2 million children annually worldwide, with the greatest burden being in Africa and Southeast Asia (5–7). Consequently, failure to perform LP when indicated can contribute to diagnostic uncertainty and delayed or inappropriate management (3).

Even in settings where LP has been well established as best practice, its use in routine care is often suboptimal. Pediatric hospitals in high-income countries have reported declining LP rates, indicating that barriers to LP are not confined to resource-limited settings (8). In Africa, however, additional patient, healthcare worker and health system level barriers may further limit access to diagnostic LP. Zambian research has identified factors influencing LP performance including healthcare worker confidence, clinical decision-making, availability of equipment and concerns surrounding the procedure (9). Qualitative work involving Zambian patients, caregivers, doctors and nurses further demonstrated that perceptions of LP, communication between health workers and families, concerns about pain and complications, and wider social and health-system factors may influence whether an LP is undertaken (10). Caregiver refusal is also a recognised barrier. Studies have reported substantial parental refusal of LP among children with suspected CNS infection and have identified fear, misconceptions and concerns about the procedure among factors contributing to refusal (11–14). In Africa, concerns surrounding LP refusal have been sufficiently recognised to prompt calls for interventions that better understand and address community and healthcare-provider perceptions of the procedure (12,14).

Interventions aimed at improving LP practice have shown that this problem can be addressed. Training-based interventions have shown that LP rates can be improved. In Malawi, a healthcare-worker training intervention raised LP rates for meningitis surveillance at secondary-level facilities (15). In addition, a multipronged intervention addressing diagnosis and early management of suspected acute brain infections across Brazil, India and Malawi demonstrated that coordinated, system-level interventions, rather than training alone, can improve clinical processes (16). These findings suggest that improving LP practice requires attention not only to healthcare worker skills and confidence but also to institutional processes, diagnostic capacity and communication with families.

There remains limited information on how often pediatric LPs are actually performed in routine care in hospitals in Africa, or on whether training-based interventions produce sustained change in busy referral settings. We sought to evaluate this during the Short Intensified Treatment for Children with Tuberculous Meningitis (SURE) trial, the largest ever randomised controlled trial for the treatment of pediatric TBM, that recruited across sites in Asia and Africa (2).

### Context

A nested diagnostic sub-study of the SURE trial, SURE+DP, recruited participants in whom a lumbar puncture was clinically indicated in recruiting African hospitals in Kampala, Uganda; Lusaka, Zambia and Harare, Zimbabwe, and this was integrated into the pre-screening and screening procedures for the SURE trial (2,17). We identified that CSF sampling appeared infrequent when compared to the number of children presenting with syndromes compatible with CNS infection, the population size served, and the high morbidity and mortality settings.

#### A. Uganda

The study was conducted at Mulago National Referral Hospital, Kampala serving over 4 million people. Recruitment to SURE and SURE+DP started in May 2022 and stopped in early 2023 due to an Ebola outbreak and limited numbers of children with TBM. The Acute Care Unit, which manages acute pediatric presentations, managed approximately 450 pediatric cases per week during the study period. Because SURE recruitment had already ended, the LP training and questionnaires described below were not conducted at this site.

#### B. Zambia

University Teaching Hospital (UTH) is in Lusaka which has a population of over 2 million. Recruitment started in May 2022 and ended in July 2024. The pediatric outpatient department, which includes both the emergency room and non-acute patients, managed 750 cases per week during the study period. LP training was conducted in July 2023 (as described below) for doctors, nurses and allied health staff. We used routinely coded emergency department diagnoses to estimate how many children presented with conditions for which LP might be indicated (tuberculous meningitis, cerebral malaria, meningitis, encephalitis, hydrocephalus, other convulsions, space-occupying lesions and other CNS disorders).

#### C. Zimbabwe

Parirenyatwa and Sally Mugabe Hospitals are in Harare, with a population of over 1.5 million. The study period was identical to Zambia. In 2025 the emergency department of Parirenyatwa and Sally Mugabe Hospitals managed approximately 150 and 700 pediatric cases per week, respectively.

Emergency department attendances for the study period were not available. LP training was done in July 2023 (Parirenyatwa Hospital) and August 2023 (Sally Mugabe Hospital).

### Pediatric LP monitoring

We quantified the frequency of pediatric CSF samples received for microbiological analysis at urban tertiary hospitals that were recruiting to the SURE trial and its diagnostic sub-study SURE+DP (2,17). Recruitment took place at four urban tertiary/referral hospitals in Kampala (Uganda), Lusaka (Zambia) and Harare (Zimbabwe). As part of routine screening, weekly counts of pediatric CSF samples (from children aged 28 days to 18 years) received for microbiological analysis at the hospital microbiology laboratory or sent to external accredited laboratories as part of the study were reviewed to understand how frequently LP was being performed. Neonates were excluded. Neurosurgical cases could only be excluded in Zimbabwe.

### Family-facing video development

To support consent conversations with families, <u>an educational video</u> for caregivers was also developed. Consultation with clinical practitioners from all three participating centres and the wider SURE team, with experience in other African settings informed the narrative and key points. A pilot version was tested by healthcare practitioners and the video shortened in response, prior to translation into local languages <u>Shona</u>, <u>Bemba</u>, <u>Nyanja</u> and <u>Luganda</u>. The video was then circulated to all healthcare workers in settings where LP was performed, as a tool to aid LP consent discussions.

### Healthcare worker training

In Zambia and Zimbabwe, healthcare worker training was conducted at sites where SURE recruitment was ongoing between June and August 2023. Participants included doctors, nurses and allied health professionals. Each training session lasted approximately 4 hours and focused on the recognition, investigation and management of suspected CNS infections. Practical simulation using mannequins was incorporated to demonstrate LP technique. Knowledge, confidence, prior experience and perceived barriers to performing LP were assessed using questionnaires administered before and after training.

At UTH, Zambia, where denominator data were available, an independent two-group, two-tailed proportion test compared the ratio of CSF samples received to children with a coded diagnosis compatible with the need to consider LP, in the six months before versus after training. The diagnoses included were TB meningitis, cerebral malaria, meningitis, encephalitis, hydrocephalus, febrile convulsions, other convulsions, space-occupying lesions and other CNS disorders.

Equivalent statistical comparison was not possible in Zimbabwe, where denominator data were unavailable, or in Uganda, where recruitment (and therefore data collection) had already ended before training began.

## Findings

Median weekly pediatric CSF samples were 1 at Parirenyatwa and Sally Mugabe Hospitals, 6 at UTH and 4 at Mulago Hospital (Figure 1A-D).

**Figure 1A:**
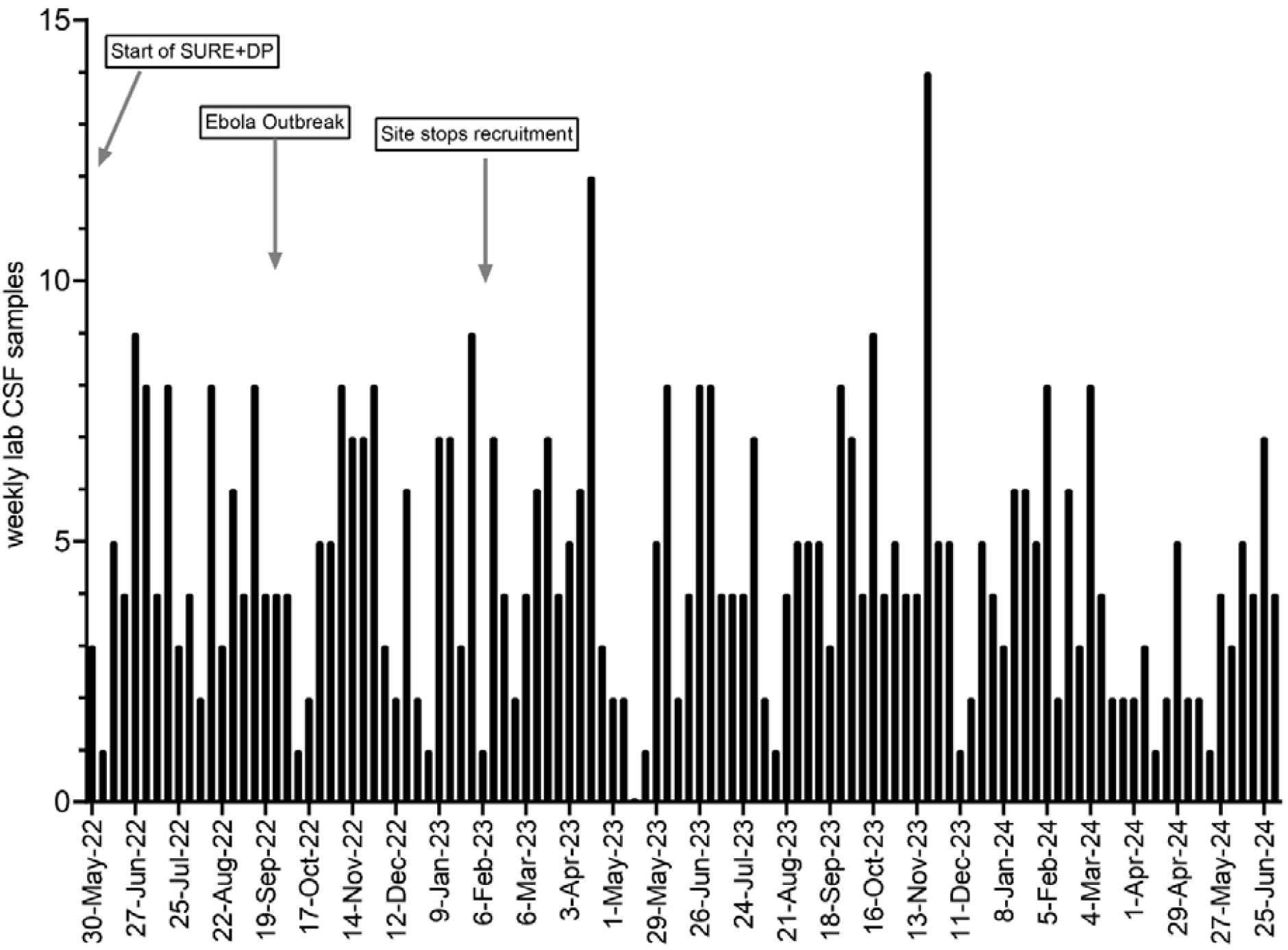
Graph of CSF samples collected per week at Mulago, Uganda from June 2022 to July 2024

**Figure 1B:**
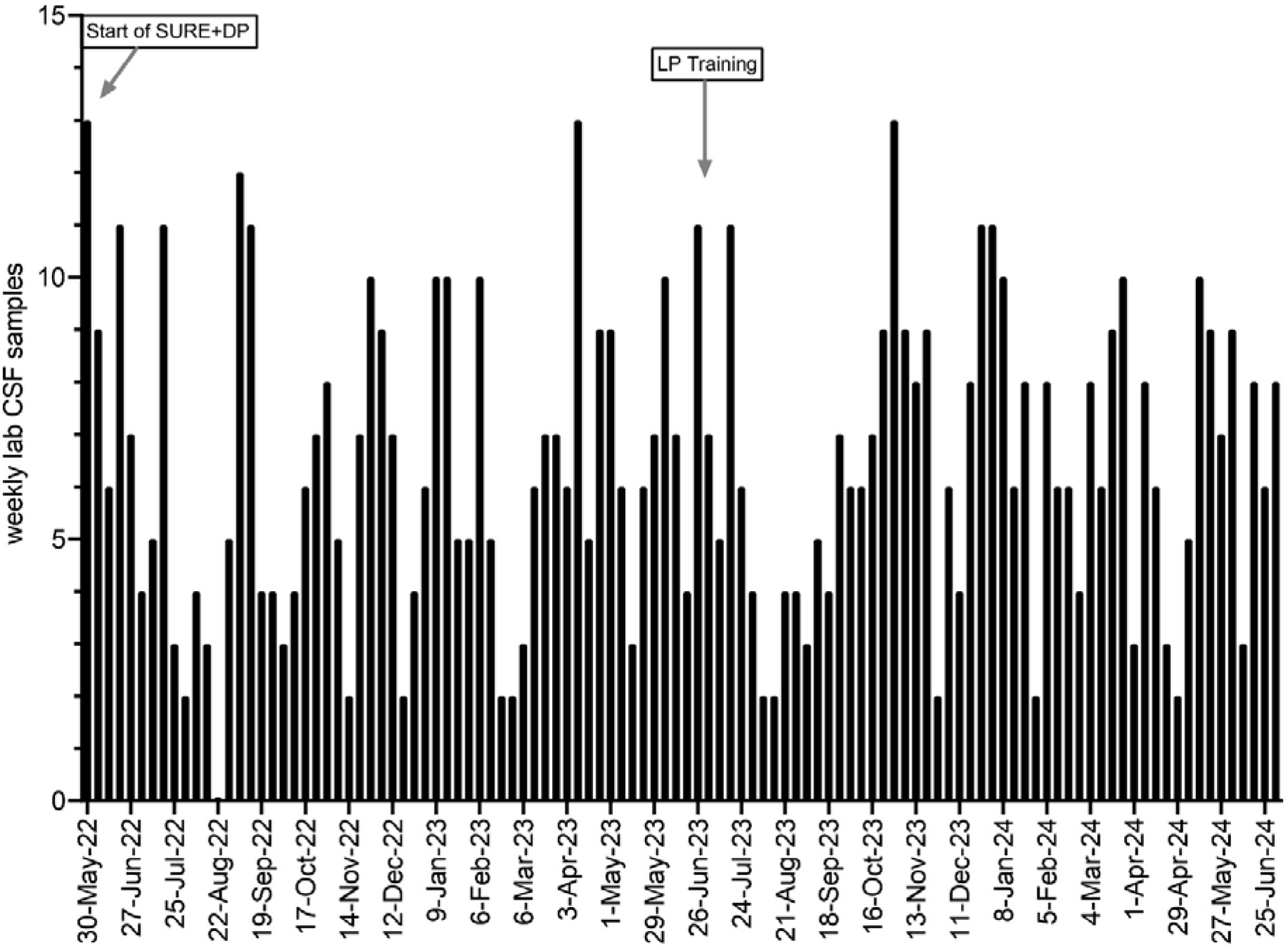
Graph of CSF samples collected per week at UTH, Zambia from June 2022 to July 2024

Overall, training significantly improved healthcare worker-reported confidence to assist and hold a patient during an LP (p<0.001 95% CI [-1.38, -0.39]) and confidence to perform LPs in children of all age groups (p<0.001). In Zimbabwe, an increase in LP procedures was observed the week immediately post-training but was not sustained (figure 1C and 1D). At UTH in Zambia, 919 children had a coded diagnosis compatible with the need to consider an LP while 705 CSF samples were received by the lab during the same period (77%). The proportion of children who had diagnoses compatible with the need to consider an LP and number of CSF samples increased from 91.6% in the 6 months before training to 98.8% in the 6 months after training (p=0.002).

**Figure 1C:**
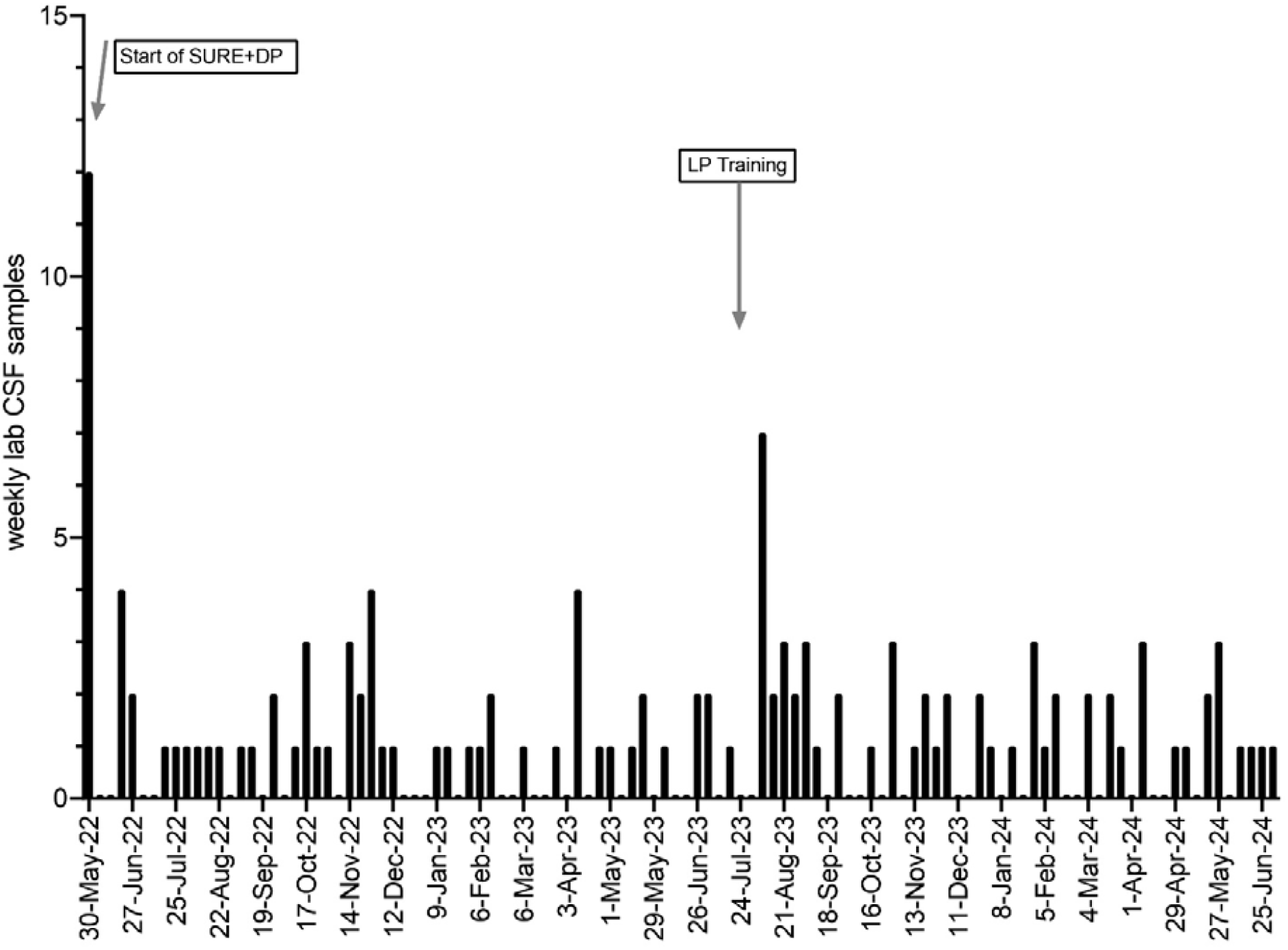
Graph of CSF samples collected per week at Sally Mugabe Hospital, Zimbabwe from June 2022 to July 2024

**Figure 1D:**
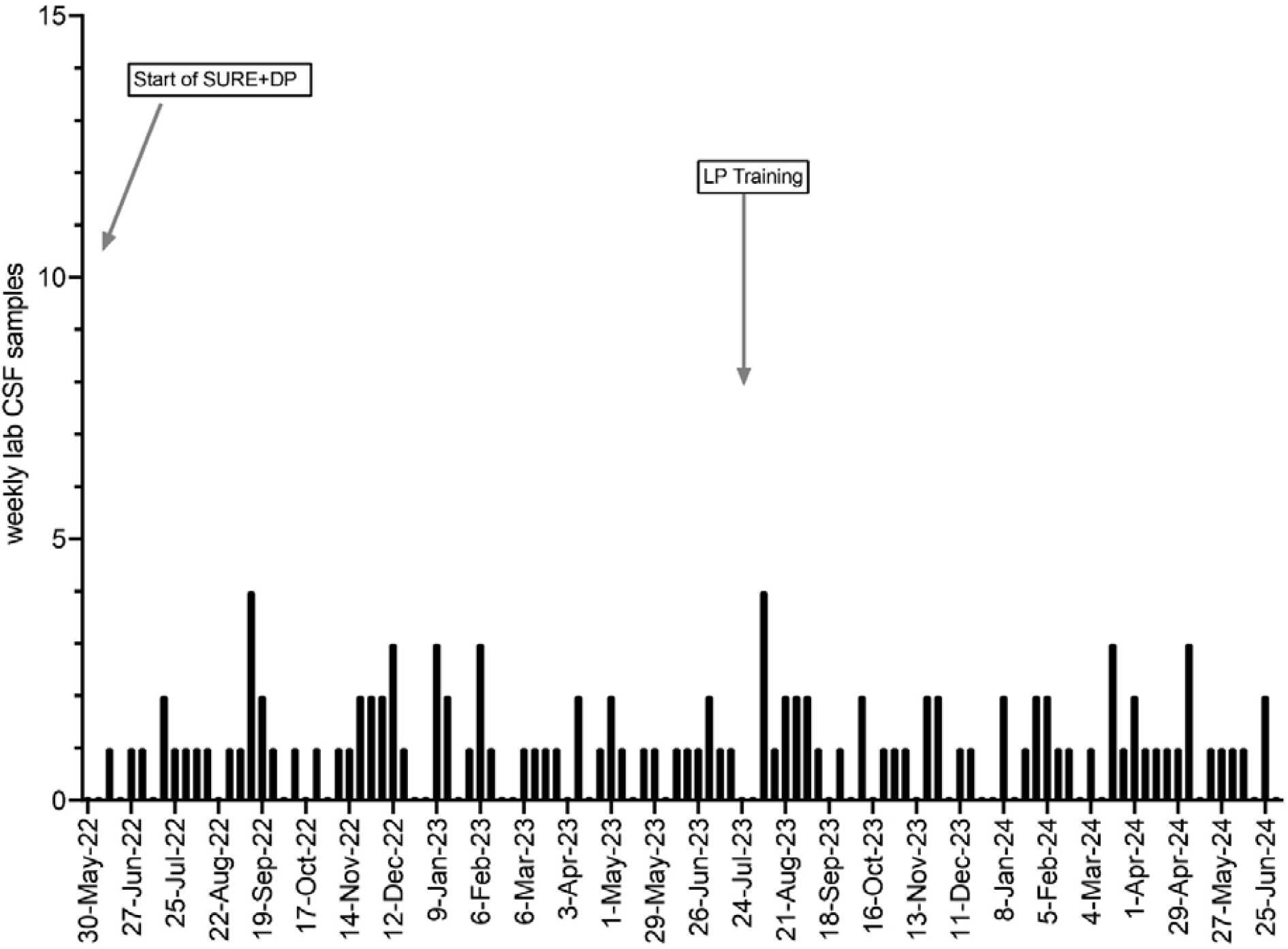
Graph of CSF samples collected per week at Parirenyatwa Hospital, Zimbabwe from June 2022 to July 2024

Healthcare workers reported that parental refusal, lack of confidence among junior staff, delays in obtaining consent from male family decision makers, and concerns over complications or resource availability such as LP needles were the main barriers to performing LP in pediatric patients (Figure 2).

**Figure 2.**
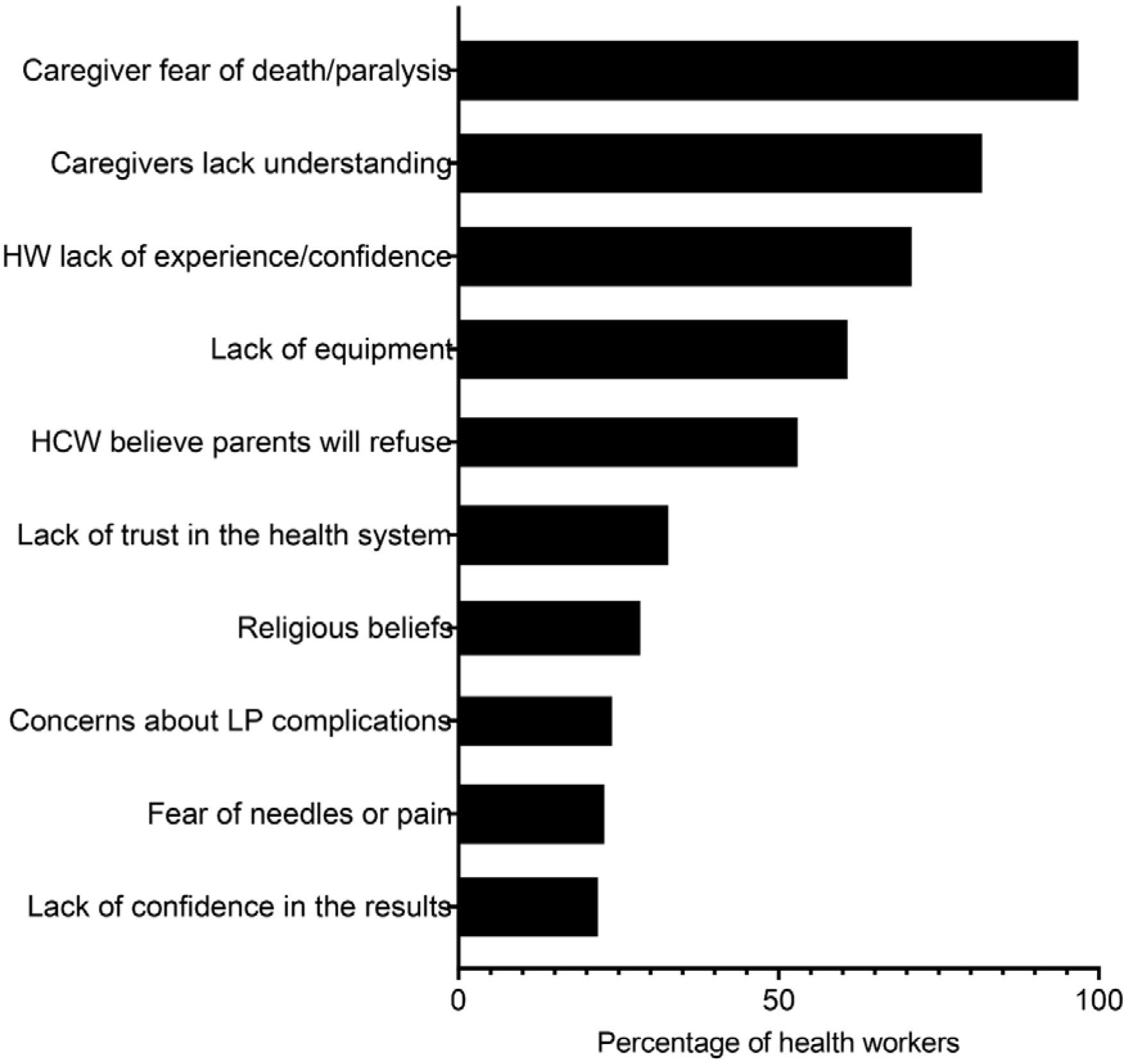
Healthcare workers perceived reasons for low LP completion in Zambia (n=18) and Zimbabwe (n=61) (total 79)

## Discussion

Across all four hospitals, pediatric LP was performed infrequently when compared to the number of children presenting with clinical features for which LP might be indicated, even though these were tertiary referral centres actively recruiting to a TBM trial. This was true regardless of country, hospital size, or whether coded diagnostic denominator data were available, suggesting the pattern reflects something structural about how LP is used in routine care rather than a feature of any single site.

There were, however, obvious differences between settings. Weekly CSF sampling at UTH and Mulago Hospital, while still low relative to the number of pediatric attendances, was higher than at the two Harare hospitals, where the median number of CSF samples was one for the study period. Several factors could have contributed to this difference. Denominator data on emergency department coded diagnoses compatible with LP were not available in Zimbabwe, so it is not possible to state with confidence what proportion of children who might have benefited from LP actually received one. A second possibility is that CSF samples reaching the laboratory in Zimbabwe may under-represent LPs performed for indications outside the trial. Nonetheless, the consistency of a low median across two hospitals over an extended period suggests that access to LP, whether relating to equipment, staffing, caregiver consent, or institutional practice, was more constrained in Harare than at the other sites during this period, and this warrants further local investigation. Thirdly, failure to exclude neurosurgical cases in Zambia could have inflated the number of CSF samples reaching the laboratory since these patients typically undergo repeated CSF sampling.

Training improved healthcare-worker confidence at both sites, but this was not consistently followed by a sustained increase in LP activity, supporting the evidence from both Malawi and the multi-country intervention in Brazil, India and Malawi, that system-level or multifaceted approaches, rather than isolated training, are more likely to produce long lasting change in diagnostic practice for suspected central nervous system infections (15,16).

The barriers reported by healthcare workers, particularly parental refusal and delays obtaining consent from male family decision makers, are consistent with earlier qualitative work in the region, and point to the need for caregiver and community-facing interventions, such as the educational video introduced alongside training in this case study, as a plausible complement to healthcare-worker training (9,10).

This community case study has several limitations. Denominator data allowing calculation of LP rates was based on coded diagnoses and were only available at one of four hospitals. The post-training observation period was relatively short, there was no concurrent control site, and confidence was self-reported rather than directly observed. Neurosurgical cases could only be excluded from CSF counts in Zimbabwe, which may affect comparability between sites. CSF samples sent to external laboratories at family and/or clinician discretion outside of the SURE and SURE+DP studies would not be captured.

## Data Availability

All data produced in the present study are available upon reasonable request to the authors

## Lessons learned and recommendation

Our findings confirm that pediatric LP rates are low in high-burden settings, even at referral hospitals participating in clinical trials. Addressing low LP rates is critical for timely diagnosis and management of CNS infections, especially given high morbidity and mortality.

Training improved healthcare worker confidence but had limited sustained impact on LP rates. Embedding routine, low-burden surveillance of CSF sampling, for example via existing trial or laboratory data systems, alongside coded clinical denominator data would allow hospitals to track LP practice over time and identify where gains are, or are not, being sustained. Caregiver-facing tools that address specific, previously identified barriers to consent, including the role of male family decision makers, may be a useful complement to workforce training and warrant formal evaluation. WHO meningitis 2025 guidelines and Brain Infections Global Health Network resources provide practical guidance that could support this type of system level strengthening (4,16,18). Additional strategies include mentorship, repeated training, institutional support, improved supply chains and community engagement to address caregiver concerns. Finally, replicating this kind of CSF surveillance across additional sites, alongside consistent collection of denominator data, would help distinguish genuinely low LP performance from gaps in how CSF sampling or coded diagnoses are captured.

Even within large referral/tertiary hospitals actively engaged in a large TBM trial, pediatric LP was performed far less often than the burden of CNS infection would suggest is needed, and training alone had limited durable effect. Efforts to close this gap most likely require combining healthcare-worker training with system-level support, caregiver engagement, and routine surveillance of LP practice.

## Author Contributions

CM conceptualised and designed the study, drafted the initial manuscript, critically reviewed and revised the manuscript and analysed the participant questionnaires from the LP training sessions. CZ conceptualised and designed the study, drafted the initial manuscript, critically reviewed and revised the manuscript and analysed the participant questionnaires from the LP training sessions. SAS conceptualised and designed the study analysed the participant questionnaires from the LP training sessions.

RB conceptualised and designed the study, drafted the initial manuscript, critically reviewed and revised the manuscript

TM, WM, DM, MK, SBK, EW, CM and CZ collected data and, critically reviewed and revised the manuscript

MS, MCS, and EF analysed the participant questionnaires from the LP training sessions

All authors critically reviewed, revised and approved the final manuscript as submitted and agreed to be accountable for all aspects of the work

## Funding/Support

The SURE trial and SURE+DP diagnostic sub-study were funded by:

Joint Global Health Trials Grant reference: MR/R006113/1 funded by Department of Health and Social Care (DHSC), the Foreign, Commonwealth & Development Office (FCDO), the Medical Research Council (MRC) and Wellcome Trust. This UK-funded award is part of the EDCTP2 programme supported by the European Union.

MRC Respiratory Programme grant MC_UU_00004/04

Meningitis Now – “SURE+DP: improving Diagnosis and Prognosis for pediatric tuberculous meningitis through the SURE treatment trial”

Medical Research UK Dorothy Temple Cross Tuberculosis International Collaboration Grant: MRF-131-0003-RG-BASU-C0851.

RB was funded by an NIHR Academic Clinical Lectureship 2019-2023 (CL-2018-20-001). SAS was funded by Nagasaki University WISE Doctoral Program for World-leading Innovative and Smart Education” for Global Health.

